# Predictive value of isolated symptoms for diagnosis of SARS-CoV-2 infection in children tested during peak circulation of the delta variant

**DOI:** 10.1101/2021.12.17.21267993

**Authors:** Adrianna L. Westbrook, Laura C. Benedit, Jennifer K Frediani, Mark A. Griffiths, Nabeel Y. Khan, Joshua M. Levy, Claudia R. Morris, Christina A. Rostad, Cheryl L. Stone, Julie Sullivan, Miriam B. Vos, Jean Welsh, Anna Wood, Greg S. Martin, Wilbur Lam, Nira R. Pollock

**Author notes:** <u>Corresponding author</u>: Nira R. Pollock, M.D., Ph.D., D(ABMM), Associate Medical Director, Infectious Diseases Diagnostic Laboratory, Boston Children’s Hospital, Division of Infectious Diseases, Beth Israel Deaconess Medical Center, Associate Professor of Pathology and Medicine, Harvard Medical School, Farley Building 8th floor, Room FA828, 300 Longwood Ave, Boston, MA, USA 02115. <u>Alternate Corresponding:</u> Wilbur A. Lam, MD, PhD, Professor and W. Paul Bowers Research Chair, Department of Pediatrics, Wallace H. Coulter Department of Biomedical Engineering, Aflac Cancer and Blood Disorders Center of Children’s Healthcare of Atlanta, Chief Innovation Officer, Pediatric Technology Center, Emory University and Georgia Institute of Technology, 2015 Uppergate Drive, Atlanta, GA 30322.

## Abstract

**Background:** COVID-19 testing policies for symptomatic children attending U.S. schools or daycare vary, and whether isolated symptoms should prompt testing is unclear. We evaluated children presenting for SARS-CoV-2 testing to determine if the likelihood of having a positive SARS-CoV-2 test differed between participants with one versus ≥2 symptoms, and to examine the predictive capability of isolated symptoms.

**Methods:** Participants ≤ 18 years presenting for clinical SARS-CoV-2 molecular testing in six sites in urban/suburban/rural Georgia (July-October, 2021; delta variant predominant) were queried about individual symptoms. Participants were classified into three groups: asymptomatic, one symptom only, or ≥2 symptoms. SARS-CoV-2 test results and clinical characteristics of the three groups were compared. Sensitivity, specificity, and positive/negative predictive values (PPV/NPV) for isolated symptoms were calculated by fitting a saturated Poisson model.

**Results:** Of 602 participants, 21.8% tested positive and 48.7% had a known or suspected close contact. Children reporting one symptom (n=82; OR=6.00, 95% CI: 2.70-13.33) and children reporting ≥2 symptoms (n=365; OR=5.25: 2.66-10.38) were significantly more likely to have a positive COVID-19 test than asymptomatic children (n=155), but they were not significantly different from each other (OR=0.88: 0.52-1.49). Sensitivity/PPV were highest for isolated fever (33%/57%), cough (25%/32%), and sore throat (21%/45%); headache had low sensitivity (8%) but higher PPV (33%). Sensitivity/PPV of isolated congestion/rhinorrhea were 8%/9%.

**Conclusions:** With high delta variant prevalence, children with isolated symptoms were as likely as those with multiple symptoms to test positive for COVID-19. Isolated fever, cough, sore throat, or headache, but not congestion/rhinorrhea, offered highest predictive value.

**Key points:** In an area with high community prevalence of the delta variant, children presenting with one symptom were as likely as those with two or more symptoms to test positive for SARS-CoV-2 infection. Isolated symptoms should be considered in testing decisions.

## Introduction

Despite the surge of pediatric SARS-CoV-2 infections due to the delta variant [1, 2] and the coincident reopening of schools, little detail exists regarding the early symptom profile for children infected with the delta variant. This lack of information complicates development of school policy regarding which symptoms, either in isolation or in combination, should potentially require exclusion of a newly symptomatic child from school or childcare facilities until SARS-CoV-2 infection has been ruled out by testing. Some policies have specifically stated that children with a single isolated symptom (e.g. headache, sore throat, congestion/rhinorrhea, fatigue) can attend school and do not need testing for SARS-CoV-2 infection (e.g. [3-5]). While a recent preprint from the UK [6] described the most common symptoms observed over the first week of illness in children with the delta (vs alpha) variant, the study did not provide information on the prevalence or predictive value of isolated symptoms, nor on the time course of symptoms.

Given that isolated symptoms are relevant for real-time testing decisions made by parents/guardians and school staff, it is important to define the relative pre-test probability of having COVID-19 for children with isolated symptoms, allowing testing policy to be refined to avoid either missing cases or unnecessarily excluding children with isolated symptoms from school. In this study, we compared rates of COVID-19 in children presenting for testing in a high prevalence area during peak circulation of the SARS-CoV-2 delta variant who reported no symptoms, one isolated symptom, or two or more symptoms. Additionally, we evaluated the predictive value of each isolated symptom and the impact of age, close contacts, and vaccination status on the likelihood of having COVID-19.

## Methods

Patients aged 0 to 18 years old presenting to one of six ambulatory testing sites in Georgia (two urban and three suburban sites in the Atlanta area and one rural site in Blairsville) of the Atlanta Center of Microsystems Engineered Point-of-Care Technologies, the test verification center of the NIH-funded Rapid Acceleration of Diagnostics (RADx) Initiative, between July 4^th^ and October 15^th^, 2021 were prospectively enrolled following informed consent and assent (as applicable per age) and participated in the following procedures: clinical nasopharyngeal PCR testing for SARS-CoV-2, detailed review of symptoms present at the time of clinical testing and overall symptom duration, and collection of additional samples for future research testing under the RADx program [7, 8]. All RADx testing sites are open to all pediatric patients and are located throughout the surrounding area as drive-through or walk-up sites (typically embedded in an existing clinical infrastructure, e.g. ambulatory urgent care clinic). There were no costs to the participants for SARS-CoV-2 testing, and all clinical test results were provided to the parents/guardians. The study time window was set to allow focus on the SARS-CoV-2 delta variant: as of the week of July 4, the delta variant comprised 78% of available sequences in US Department of Health and Human Services (HHS) Region 4 (which includes Georgia) based on the Centers for Disease Control and Prevention (CDC) SARS-CoV-2 variant surveillance program [9]. Participants and their families were asked about the presence or absence of each of the following symptoms: fever (either subjective or temperature ≥ 100.4°F accepted), chills, congestion/rhinorrhea, cough, headache, sore throat, fatigue, arthralgias, myalgias, photophobia, vomiting, nausea, diarrhea, abdominal pain, loss of sense of taste or smell, shortness of breath, or any other symptoms. Participants were categorized as being asymptomatic, reporting one symptom, or reporting two or more symptoms. Vaccination status and any known or suspected recent close contact were ascertained. This study was approved by the Institutional Review Boards at Emory University (STUDY00000932) and Children’s Healthcare of Atlanta (IRB#00001082).

### Statistical Analysis

Categorical data are displayed as N (%) while continuous variables are displayed as median (IQR). To statistically compare categorical variables, a chi-square or Fisher’s test were used. Continuous variables were compared by a Wilcoxon Rank Sum test. Odds ratios and corresponding confidence intervals were calculated using the LOGISTIC procedure. Odds ratios were also adjusted for exposure status to address potential confounding. Diagnostic accuracy measures such as sensitivity, specificity, and positive/negative predictive value were calculated using the NLMIXED procedure and a saturated Poisson model. Sensitivity analyses were conducted removing participants who had a fever or who were vaccinated. Furthermore, positivity rates among symptom groups were stratified by exposure status. All confidence intervals are 95% confidence intervals. All analyses were conducted using SAS 9.4 (Cary, NC). Graphs were created with RStudio (Boston, MA) and the ggplot2 package ([10] New York, NY).

## Results

602 pediatric patients [median age 9 years (IQR: 5-13)] were included in this analysis, including 155 asymptomatic children (25.2% previously vaccinated), 82 children who reported one symptom at the time of testing (13.4% previously vaccinated), and 365 who reported two or more symptoms at the time of testing (9.0% previously vaccinated) (**Table 1**). There were no significant differences between these three groups in terms of age, sex, race, or ethnicity. In those with only one symptom at presentation (n = 82), congestion/rhinorrhea was the most frequently reported symptom (26.8%), followed by cough (23.2%), fever (17.1%), sore throat (13.4%), and headache (7.3%). The group with one symptom and the group with two or more symptoms had equal median duration of symptoms at the time of testing [2 days (IQR 1-4)].

**Table 1.** Participant Characteristics at Time of Enrollment in RADx between July 4^th^ and October 15^th^, 2021

| Variable | Level | Overall<br>N=602 | Not<br>Symptomatic<br>N=155 | 1<br>Symptom<br>N=82 | 2+<br>Symptoms<br>N=365 | P-value* |
| --- | --- | --- | --- | --- | --- | --- |
| Enrollment<br>location | Blairsville<sup>1</sup> | 130 (21.6%) | 40 (25.8%) | 17<br>(20.7%) | 73 (20.0%) | 0.003 |
|  | Hospital – Egleston<sup>2</sup> | 63 (10.5%) | 1 (0.7%) | 10<br>(12.2%) | 52 (14.3%) |  |
|  | Urgent Care – Satellite<br>Boulevard<sup>3</sup> | 175 (29.1%) | 12 (7.7%) | 18<br>(22.0%) | 145 (39.7%) |  |
|  | Urgent Care - Town<br>Center<sup>4</sup> | 61 (10.1%) | 5 (3.2%) | 12<br>(14.6%) | 44 (12.1%) |  |
|  | Viral Solutions – Atlanta<br>Public Schools<sup>5</sup> | 15 (2.5%) | 3 (1.9%) | 3 (3.7%) | 9 (2.5%) |  |
|  | Viral Solutions –<br>Decatur<sup>6</sup> | 158 (26.3%) | 94 (60.7%) | 22<br>(26.8%) | 42 (11.5%) |  |
| Age | Median (IQR) | 9.0 (5.0-13.0) | 10.0 (7.0-14.0) | 9.0 (6.0-13.0) | 9.0 (5.0-12.0) | 0.50 |
| Sex | Female | 284 (47.2%) | 78 (50.3%) | 35<br>(42.7%) | 171 (46.9%) | 0.49 |
|  | Male | 318 (52.8%) | 77 (49.7%) | 47<br>(57.3%) | 194 (53.2%) |  |
| Race | Asian | 14 (2.3%) | 3 (1.9%) | 5 (6.1%) | 6 (1.6%) | 0.11 |
|  | Black/African American | 193 (32.1%) | 28 (18.1%) | 28<br>(34.2%) | 137 (37.5%) |  |
|  | Other | 60 (10.0%) | 13 (8.4%) | 6 (7.3%) | 41 (11.2%) |  |
|  | White | 324 (53.8%) | 110 (71.0%) | 40<br>(48.8%) | 174 (47.7%) |  |
|  | Declined to<br>Answer/Missing | 11 (1.8%) | 1 (0.7%) | 3 (3.7%) | 7 (1.9%) |  |
| Ethnicity | Hispanic | 81 (13.5%) | 13 (8.4%) | 12<br>(14.6%) | 56 (15.3%) | 0.62 |
|  | Non-Hispanic | 515 (85.5%) | 139 (89.7%) | 69<br>(84.2%) | 307 (84.1%) |  |
|  | Declined to<br>Answer/Missing | 6 (1.0%) | 3 (1.9%) | 1 (1.2%) | 2 (0.6%) |  |
| Symptoms | Fever | 221 (36.7%) | 0 (0.0%) | 14<br>(17.1%) | 207 (56.7%) | <0.001 |
|  | Chills | 44 (7.3%) | 0 (0.0%) | 0 (0.0%) | 44 (12.1%) | <0.001 |
|  | Congestion/rhinorrhea | 279 (46.4%) | 0 (0.0%) | 22<br>(26.8%) | 257 (70.4%) | <0.001 |
|  | Cough | 270 (44.9%) | 0 (0.0%) | 19<br>(23.2%) | 251 (68.8%) | <0.001 |
|  | Headache | 130 (21.6%) | 0 (0.0%) | 6 (7.3%) | 124 (34%) | <0.001 |
|  | Sore throat | 162 (26.9%) | 0 (0.0%) | 11<br>(13.4%) | 151 (41.4%) | <0.001 |

| Variable | Level | Overall<br>N=602 | Not<br>Symptomatic<br>N=155 | 1<br>Symptom<br>N=82 | 2+<br>Symptoms<br>N=365 | P-value* |
| --- | --- | --- | --- | --- | --- | --- |
|  | Fatigue | 49 (8.1%) | 0 (0.0%) | 0 (0.0%) | 49 (13.4%) | <0.001 |
|  | Arthralgias | 4 (0.7%) | 0 (0.0%) | 0 (0.0%) | 4 (1.1%) | 1.00 |
|  | Myalgias | 27 (4.5%) | 0 (0.0%) | 0 (0.0%) | 27 (7.4%) | 0.01 |
|  | Photophobia | 2 (0.3%) | 0 (0.0%) | 0 (0.0%) | 2 (0.5%) | 1.00 |
|  | Vomiting | 47 (7.8%) | 0 (0.0%) | 3 (3.7%) | 44 (12.1%) | 0.03 |
|  | Nausea | 33 (5.5%) | 0 (0.0%) | 0 (0.0%) | 33 (9%) | 0.005 |
|  | Diarrhea | 40 (6.6%) | 0 (0.0%) | 3 (3.7%) | 37 (10.1%) | 0.06 |
|  | Abdominal pain | 42 (7.0%) | 0 (0.0%) | 3 (3.7%) | 39 (10.7%) | 0.049 |
|  | Loss of sense of taste or<br>smell | 22 (3.7%) | 0 (0.0%) | 1 (1.2%) | 21 (5.8%) | 0.10 |
|  | Shortness of breath | 16 (2.7%) | 0 (0.0%) | 0 (0.0%) | 16 (4.4%) | 0.05 |
|  | Other symptom(s) | 15 (2.5%) | 0 (0.0%) | 0 (0.0%) | 14 (3.8%) | 0.08 |
| Duration of<br>Symptoms at<br>Time of Test | Median (IQR) | 2.0 (1.0-4.0)** | -- | 2.0 (1.0-4.0) | 2.0 (1.0-4.0) | 0.17 |
| Received COVID<br>vaccine | I don't know | 4 (0.7%) | 1 (0.7%) | 0 (0.0%) | 3 (0.8%) | 0.36 |
|  | No | 514 (85.4%) | 114 (73.6%) | 71<br>(86.6%) | 329 (90.1%) |  |
|  | Yes | 83 (13.8%) | 39 (25.2%) | 11<br>(13.4%) | 33 (9.0%) |  |
|  | Missing | 1 (0.2%) | 1 (0.7%) | 0 (0.0%) | 0 (0.0%) |  |
| Exposure status | Known contact | 238 (39.5%) | 68 (43.9%) | 40<br>(48.8%) | 130 (35.6%) | 0.08 |
|  | Suspected contact only | 55 (9.1%) | 16 (8.5%) | 7 (8.5%) | 32 (8.8%) |  |
|  | Neither known nor<br>suspected contacts | 309 (51.3%) | 71 (45.8%) | 35<br>(42.7%) | 203 (55.6%) |  |
| Exposure status | Known or suspected<br>contact | 293 (48.7%) | 84 (54.2%) | 47<br>(57.3%) | 162 (44.4%) | 0.04 |
|  | No known or suspected<br>contact | 309 (51.3%) | 71 (45.8%) | 35<br>(42.7%) | 203 (55.6%) |  |

1) Rural, outdoor testing event; 2) Urban, city of Atlanta Emergency Department; 3) Suburban, urgent care clinic; 4) Suburban, urgent care clinic; 5) Urban, dedicated testing site; 6) Suburban, dedicated testing site
\*p-values are for the statistical comparison between 1 symptom and 2+ symptom groups.
\*\*Those with no symptoms were excluded from this calculation

Overall, 48.7% of participants had a known and/or suspected close contact with COVID-19. Those with one symptom had a significantly higher proportion of exposure to a known and/or suspected contact with COVID-19 than those with two or more symptoms (57.3% vs 44.4%, p=0.04; **Table 1**); however, those with one symptom and those with no symptoms had a similar proportion of exposure (57.3% vs 54.2%, p=0.65).

Of the 602 patients, 21.8% tested positive by PCR (6.5% of asymptomatic children, 29.3% of children with only one symptom, and 26.6% of children with two or more symptoms; **Table 2**). Children with only one symptom were 6.00 (95% CI: 2.70, 13.33) times as likely to test positive for COVID-19 as children with no symptoms. Children with two or more symptoms were 5.25 (95% CI: 2.66, 10.38) times as likely to test positive for COVID-19 as children with no symptoms. Interestingly, children with two or more symptoms were not statistically more likely to test positive for COVID-19 compared to children with only 1 symptom (OR=0.88, 95% CI: 0.52, 1.49) indicating that the likelihood of having COVID-19 was not dependent on the number of symptoms, but rather that a symptom was present (**Table 3**). These findings remained true when children with fever (**Table 2**), vaccinated children (**Supplementary Table 1**), or those with both fever/vaccination (**Supplementary Table 1**) were excluded from analysis. Additionally, these findings remained consistent after controlling for exposure status (**Table 3, Table 4**). Holding known or suspected exposure status constant, those with one symptom (OR=6.20, 95% CI: 2.76-13.97) and those with two or more symptoms (OR=6.29, 95% CI: 3.14-12.60) had significantly higher odds of having a positive COVID-19 test compared to those with no symptoms, but these groups did not significantly differ from each other (OR=1.02, 95% CI: 0.59-1.76) (**Table 3, Table 4**). These findings remained true when vaccinated children were excluded (**Supplementary Table 2**).

**Table 2.** Percentage of Children Who Tested Positive by Symptom Status

| Symptom Status | All Participants |  | Excluding Participants with Fever |  |
| --- | --- | --- | --- | --- |
|  | Total Participants | Participants Who Tested Positive (%) | Total Participants | Participants Who Tested Positive (%) |
| 0 symptoms | 155 | 10 (6.5%) | 155 | 10 (6.5%) |
| 1 symptom | 82 | 24 (29.3%) | 68 | 16 (23.5%) |
| 2+ symptoms | 365 | 97 (26.6%) | 158 | 30 (19.0%) |
|  | Vaccinated Participants |  | Vaccinated Participants without Fever |  |
|  | Total Participants | Participants Who Tested Positive (%) | Total Participants | Participants Who Tested Positive (%) |
| 0 symptoms | 39 | 0 (0.0%) | 39 | 0 (0.0%) |
| 1 symptom | 11 | 1 (9.1%) | 10 | 1 (10.0%) |
| 2+ symptoms | 33 | 3 (9.1%) | 21 | 1 (4.8%) |

**Table 3.** Logistic Regression Analysis Modeling Association between Number of Symptoms and COVID-19 Test Positivity in the Overall Population and By School Age Groups

|  | Overall, N=602 |  | School Age Groups |  |  |  |  |  |
| --- | --- | --- | --- | --- | --- | --- | --- | --- |
|  |  |  | Elementary School<br>(≤10 y), N=353 |  | Middle School<br>(11-13 y), N=111 |  | High School<br>(≥ 14 y), N=138 |  |
|  | OR<br>(95% CI) | P | OR<br>(95% CI) | P | OR<br>(95% CI) | P | OR<br>(95% CI) | P |
| *Unadjusted odds ratios* |  |  |  |  |  |  |  |  |
| 1 vs 0 Symptoms | 6.00<br>(2.70, 13.33) | <0.001 | 4.06<br>(1.58, 10.39) | 0.004 | 19.43<br>(1.88, 201.17) | 0.01 | 10.00<br>(1.04, 96.47) | 0.047 |
| 2+ vs 0 Symptoms | 5.25<br>(2.66, 10.38) | <0.001 | 1.99<br>(0.89, 4.48) | 0.09 | 21.25<br>(2.74, 165.13) | 0.004 | 26.96<br>(3.52, 206.45) | 0.002 |
| 2+ vs 1 Symptom | 0.88 (0.52, 1.49) | 0.62 | 0.49<br>(0.25, 0.97) | 0.04 | 1.09 (0.29, 4.12) | 0.89 | 2.70 (0.82, 8.83) | 0.10 |
| *Odds ratios adjusted for exposure status (known and/or suspected contact vs no contact)* |  |  |  |  |  |  |  |  |
| 1 vs 0 Symptoms | 6.20<br>(2.76, 13.97) | <0.001 | 4.58<br>(1.73, 12.11) | 0.002 | 16.39<br>(1.56, 172.45) | 0.02 | 10.47<br>(1.08, 101.88) | 0.04 |
| 2+ vs 0 Symptoms | 6.29<br>(3.14, 12.60) | <0.001 | 2.69<br>(1.17, 6.19) | 0.02 | 20.54<br>(2.63, 160.45) | 0.004 | 29.66<br>(3.83, 229.51) | 0.001 |
| 2+ vs 1 Symptom | 1.02 (0.59, 1.76) | 0.96 | 0.59<br>(0.29, 1.20) | 0.15 | 1.25 (0.32, 2.85) | 0.74 | 2.83 (0.85, 9.43) | 0.09 |

OR=odds ratio, CI=confidence interval

**Table 4.** Percentage of Children Who Had a Known or Suspected Close Contact and Who Tested Positive by Symptom Status

| Symptom Status | Total Participants | Participants Who Had a Known or Suspected Close Contact (%) | Test Positive Rate (%) in Participants with a Known or Suspected Close Contact (%) | Participants Who Did Not Have a Known or Suspected Close Contact (%) | Test Positive Rate (%) in Participants with No Known or Suspected Close Contact |
| --- | --- | --- | --- | --- | --- |
| *All participants* |  |  |  |  |  |
| 0 symptoms | 155 | 84 (54.2%) | 9 (10.0%) | 71 (45.8%) | 1 (1.4%) |
| 1 symptom | 82 | 47 (57.3%) | 20 (42.6%) | 35 (42.7%) | 4 (11.4%) |
| 2+ symptoms | 365 | 162 (44.4%) | 61 (37.7%) | 203 (55.6%) | 36 (17.7%) |

When the cohort was broken down into three age groups that roughly mirror K-12 school organization (elementary, age 10 and under; middle school, age 11-13; high school, age 14-18), similar overall findings were observed (**Table 3, Table 5**). Elementary, middle, and high school-aged children with only one symptom were 4.06 (95% CI: 1.58, 10.39), 19.43 (95% CI: 1.88, 201.17), and 10.00 (95% CI: 1.04, 96.47) times as likely, respectively, to test positive for COVID-19 as children with no symptoms. Elementary, middle, and high school-aged children with two or more symptoms were 1.99 (95% CI: 0.89, 4.48), 21.25 (95% CI: 2.74, 165.13), and 26.96 (95% CI: 3.52, 206.45) times as likely, respectively, to test positive for COVID-19 than children with no symptoms, though this difference was not significant in elementary-aged children. Finally, elementary, middle, and high school-aged children with two or more symptoms were not more likely to test positive for COVID-19 compared to children with only one symptom (elementary school OR=0.49, 95% CI: 0.25, 0.97; middle school OR=1.09, 95% CI: 0.29, 4.12; and high school OR =2.70, 95% CI: 0.82, 8.83), indicating that likelihood of having COVID-19 was dependent only on the presence of a symptom, not on the number of symptoms present. These patterns remained similar when those who presented with fever were excluded (**Table 5**) and after adjustment for exposure status (**Table 3**).

**Table 5.** Percentage of Kids Who Tested Positive by Symptom Status and Age Group

| Symptom Status | Total<br>Participants | Participants Who Tested<br>Positive (%) | Total<br>Participants | Participants Who Tested<br>Positive (%) |
| --- | --- | --- | --- | --- |
| All Participants |  |  | Excluding Participants with Fever |  |
| Elementary Aged Children (10 and under) (N=353 in total, N=201 excluding fever) |  |  |  |  |
| 0 symptoms | 79 | 8 (10.1%) | 79 | 8 (10.1%) |
| 1 symptom | 51 | 16 (31.4%) | 39 | 9 (23.1%) |
| 2+ symptoms | 223 | 41 (18.4%) | 83 | 10 (12.1%) |
| Middle School Aged Children (11 to 13) (N=111 in total, N=77 excluding fever) |  |  |  |  |
| 0 symptoms | 35 | 1 (2.9%) | 35 | 1 (2.9%) |
| 1 symptom | 11 | 4 (36.4%) | 10 | 4 (40.0%) |
| 2+ symptoms | 65 | 25 (38.5%) | 32 | 9 (28.1%) |
| High School Aged Children (14 and over) (N=138 in total, N=103 excluding fever) |  |  |  |  |
| 0 symptoms | 41 | 1 (2.4%) | 41 | 1 (2.4%) |
| 1 symptom | 20 | 4 (20.0%) | 19 | 3 (15.8%) |
| 2+ symptoms | 77 | 31 (40.3%) | 43 | 11 (25.6%) |

Analysis of the sensitivity, specificity, and positive/negative predictive value (PPV/NPV) of isolated symptoms in this cohort is presented in **Table 6**. Notably, the PPV of isolated sore throat (45%), headache (33%), and cough (32%) were not far below the PPV of fever (57%); in contrast, the PPV of isolated congestion/rhinorrhea was 9%. The isolated symptoms with the highest sensitivity were fever (33%), cough (25%), and sore throat (21%); congestion/rhinorrhea and headache each had a sensitivity of 8%. Children who had congestion/rhinorrhea only (OR=2.47, 95% CI: 0.56-10.84) were not more likely to have a positive COVID result than children who did not have congestion/rhinorrhea. The same patterns remained true when vaccinated children were excluded (**Supplementary Table 3**).

**Table 6.** Diagnostic Accuracy of Isolated Symptoms in Children who Presented with One Symptom

| Symptom | Overall<br>N=82 | Negative<br>N=58 | Positive<br>N=24 | Sensitivity | Specificity | PPV | NPV |
| --- | --- | --- | --- | --- | --- | --- | --- |
| Fever | 14 | 6 | 8 | 0.33<br>(0.14, 0.52) | 0.90<br>(0.82, 0.97) | 0.57<br>(0.31, 0.83) | 0.76<br>(0.66, 0.87) |
| Chills | 0 | 0 | 0 | NA | NA | NA | NA |
| Congestion/<br>rhinorrhea | 22 | 20 | 2 | 0.08<br>(0.00, 0.19) | 0.66<br>(0.53, 0.78) | 0.09<br>(0.00, 0.21) | 0.63<br>(0.51, 0.76) |
| Cough | 19 | 13 | 6 | 0.25<br>(0.08, 0.42) | 0.78<br>(0.67, 0.88) | 0.32<br>(0.11, 0.53) | 0.71<br>(0.60, 0.83) |
| Headache | 6 | 4 | 2 | 0.08<br>(0.00, 0.19) | 0.93<br>(0.87, 1.00) | 0.33<br>(0.00, 0.71) | 0.71<br>(0.61, 0.81) |
| Sore throat | 11 | 6 | 5 | 0.21<br>(0.05, 0.37) | 0.90<br>(0.82, 0.97) | 0.45<br>(0.16, 0.75) | 0.73<br>(0.63, 0.84) |
| Fatigue | 0 | 0 | 0 | NA | NA | NA | NA |
| Arthralgias | 0 | 0 | 0 | NA | NA | NA | NA |
| Myalgias | 0 | 0 | 0 | NA | NA | NA | NA |
| Photophobia | 0 | 0 | 0 | NA | NA | NA | NA |
| Vomiting | 3 | 3 | 0 | 0.00<br>(0.00, 0.00) | 0.95<br>(0.89, 1.00) | 0.00<br>(0.00, 0.00) | 0.70<br>(0.60, 0.80) |
| Nausea | 0 | 0 | 0 | NA | NA | NA | NA |
| Diarrhea | 3 | 3 | 0 | 0.00<br>(0.00, 0.00) | 0.95<br>(0.89, 1.00) | 0.00<br>(0.00, 0.00) | 0.70<br>(0.60, 0.80) |
| Abdominal pain | 3 | 3 | 0 | 0.00<br>(0.00, 0.00) | 0.95<br>(0.89, 1.00) | 0.00<br>(0.00, 0.00) | 0.70<br>(0.60, 0.80) |
| Loss of sense of taste or<br>smell | 1 | 0 | 1 | 0.04<br>(0.00, 0.12) | 1.00<br>(1.00, 1.00) | 1.00<br>(1.00, 1.00) | 0.72<br>(0.62, 0.81) |
| Shortness of breath | 0 | 0 | 0 | NA | NA | NA | NA |

**Figure 1** shows variation of positive predictive value (PPV) with variation in community COVID-19 prevalence rates (ranging from 0-30%, as an estimate of peak positivity in the United States, [11]) for the isolated symptoms of congestion/rhinorrhea, cough, fever, headache, and sore throat. The overall prevalence in our cohort (21.8%) is marked. As expected, lower disease prevalence confers lower PPV for each isolated symptom, such that the most prevalent isolated symptoms do not ultimately offer high predictive capabilities.

**Figure 1:**
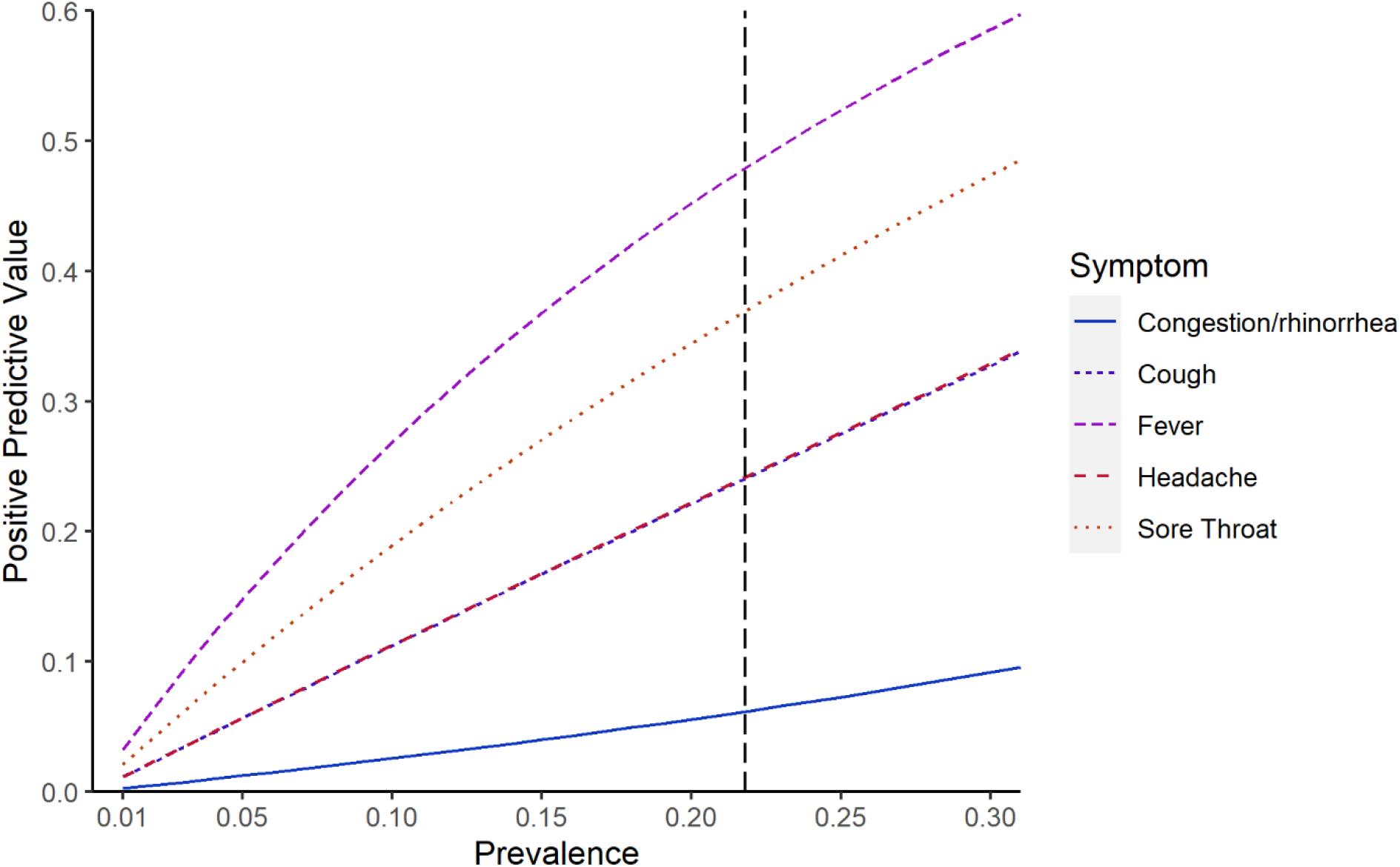
Positive Predictive Value of Isolated Symptoms Across Varying Prevalence Points This figure demonstrates the increasing positive predictive value of selected isolated symptoms across a range of prevalence points from 0% to 30%. The vertical dashed line represents the prevalence in our study cohort (21.8%).

## Discussion

In a population of children receiving ambulatory testing in an area with high community prevalence of the SARS-CoV-2 delta variant and high rates of known or suspected contact with a COVID-19 case, children presenting with one reported symptom at the time of testing were as likely as those reporting two or more symptoms to test positive for SARS-CoV-2 infection, and significantly more likely than asymptomatic children to test positive. These findings remained consistent with exclusion of children with fever or vaccination and after controlling for known/suspected close contacts in sensitivity analyses, and were true for all three school age groups (≤ 10, 11-13, and ≥ 14 years old).

The positive predictive values of isolated sore throat (45%), headache (33%), and cough (32%) were not far below the PPV of fever (57%), while the PPV of isolated congestion/rhinorrhea was only 9%. The same pattern remained true when vaccinated children were excluded. This finding would suggest that isolated sore throat, headache, and cough may need to be considered as grounds for school exclusion and COVID testing prior to return to school when community rates of COVID-19 are high. This approach would be consistent with some published recommendations: K-12 school guidance from the CDC [12] provides a list of symptoms potentially consistent with COVID-19 for which caregivers should monitor (fever, sore throat, cough, difficulty breathing, diarrhea or vomiting, or new onset of severe headache), and states that any symptom on that list should prompt testing for COVID-19. The American Academy of Pediatrics [13] provides a list of symptoms consistent with COVID-19 with the guidance that “any of the following” might be an indication for testing. However, in contrast, multiple school guidelines (including broad recommendations for K-12 schools in Massachusetts, Georgia, and California, as examples) recommend that isolated symptoms like sore throat, headache, fatigue, or rhinorrhea/congestion should not prompt testing, requiring testing only if these symptoms are present in combination with other symptoms [3-5]. Having a better understanding of the predictive value of isolated symptoms for having COVID-19 can help parents and caregivers, school nurses and administrators, and pediatricians navigate testing decisions. Moreover, identifying high likelihood symptoms could potentially aid in minimizing school-related cases and quarantines, as well as exposures of students who might be medically fragile.

This appears to be one of the first analyses of isolated symptoms present in children at the time of the actual SARS-CoV-2 test. Molteni et. al. [6] evaluated cumulative symptoms present over the entire duration of illness (potentially including *after* the actual testing was performed); a previous symptom study from the same team used a similar approach [14]. The most similar analysis in the Molteni study assessed symptom burden over the first week (<7days) of illness and found that the most common symptoms in young children (5-11 y) with the delta variant were headache (61%), rhinorrhea (54%), and fatigue (49%); in older children (12-17), the most common symptoms were headache (74%), sore throat (61%), and fatigue (60%). Notably, these symptoms and frequencies were very similar to those observed with the alpha variant, which suggests that symptom profiles with new/future variants (e.g. omicron) could also be similar. These symptoms mirror our own findings in terms of the most frequently observed symptoms in our study population, suggesting that the symptom presentations in our study cohort were generalizable. However, Molteni’s reporting of total symptom burden (number) over the first week of illness (median 4 symptoms in younger children and 6 in older children) still leaves an open question about symptoms observed earliest in the disease course. We note that in our cohort, the median duration of symptoms at the time of testing was two days (IQR one to four days) in both children with isolated symptoms and those with two or more symptoms, suggesting that our findings are likely to apply to children with new onset of isolated symptoms.

Taken together, our data and the Molteni data [6] suggest that while most children with SARS-CoV-2 ultimately will have multiple symptoms, children with new-onset isolated symptoms may need testing for SARS-CoV-2 before returning to school or childcare. Our findings would suggest that isolated symptoms to be prioritized for testing include sore throat, headache, and cough, in addition to fever (and the highly specific symptom of loss of taste and smell; we did not have sufficient data on isolated myalgias, arthralgias, fatigue, or abdominal symptoms to draw any conclusions). We note that headache had relatively high PPV but low sensitivity, and this symptom should be evaluated in a larger cohort. Given that these symptoms could also be consistent with other respiratory virus infections and strep throat, our findings in turn mean that greater access to high-sensitivity, expedited testing for these children would need to be made available to quickly exclude SARS-CoV-2 and minimize the time out of school. Congestion/rhinorrhea had lower predictive value in our study, despite its high prevalence [potential contributions to prevalence of this symptom could include other circulating viruses (e.g. rhinovirus) or allergic rhinitis with elevated pollen counts during the study period]. Decisions about school testing policy must take testing access and loss of educational continuity into account; given the low predictive value even in this high SARS-CoV-2 prevalence cohort and the high frequency with which isolated congestion/rhinorrhea occurs in school-aged children, it may not be optimal to require school exclusion and testing for those with isolated congestion/rhinorrhea.

Strengths of our study include that we prospectively collected detailed symptom data at the time of testing by interview of the patient and their family (rather than being reported virtually/on an app as in prior studies [6, 14]), making our data relevant to real-time decision-making. There were no specific symptom criteria required for testing at the study sites, allowing a relatively unbiased assessment of symptom profiles (though this does not account for testing policies that may have directed patients and families to come in for testing). Limitations include that the study was relatively small, limiting numbers for each individual isolated symptom and for subgroup analysis, and we did not have information on the severity of each symptom nor on reasons for testing in asymptomatic children. We did not include data on Ct values or viral loads in the positive samples, both due to missing data and because many different PCR assays were used for clinical testing, which would have made these data difficult to combine for analysis. We did not do sequencing to confirm that the delta variant strain was responsible for the SARS-CoV-2 infections in these children, but the time window selected is consistent with the majority having been due to this variant [9] (prior studies similarly used time windows as a proxy for variant circulation [6]). We had too few vaccinated children in this study to draw conclusions about symptom presentation in this subgroup of children (prior studies similarly included predominantly unvaccinated children [6]). The study was performed in a region with high prevalence of the delta variant and in a population with a high proportion of known or suspected close contact, with variable local testing requirements and testing availabilities, so the population who presented for testing may not be fully generalizable to other settings (including settings with lower disease/exposure prevalence or other circulating variants). Though the high exposure rates might suggest that some patients presented for testing due to exposure rather than isolated symptoms, symptom status was associated with having a positive COVID-19 test independently of exposure status. We also note that the observed prevalence in this study cohort was similar to overall school-aged prevalence in Georgia during the study time period, per the Georgia Department of Public Health (15.4%, [15]). Finally, we recognize that local circulation of other respiratory viruses would also impact SARS-CoV-2 test positivity rates and predictive values of symptoms.

In summary, our findings demonstrate that in a setting with high community prevalence of the SARS-CoV-2 delta variant, children with isolated symptoms were as likely to have COVID-19 as children with multiple symptoms. The symptoms with highest predictive value were sore throat, cough, headache, and fever; isolated congestion/rhinorrhea had high prevalence but lower predictive value. While further research is needed to understand the extent to which these findings can be generalized to other settings, these results suggest that school and daycare policies should consider isolated symptoms as potential triggers for exclusion and testing.

## Data Availability

All data produced in the present study are available upon reasonable request to the authors.

## Notes

### Author Contributions

A.L.W. and N.R.P. conceived and designed the analysis. C.L.S., L.B., N.Y.K., J.S., C.A.R., J.K.F., M.A.G., J.M. L., J.W., M.B.V., C.R.M., G.S.M., and W.L. contributed to RADx study design and implementation, including patient enrollment and data collection. A.L.W., A.W., and N.R.P. analyzed and interpreted the data. A.L.W. and N.R.P. drafted the manuscript. Critical revisions to the manuscript were made by all members of the study group. All authors had full access to all the data in the study and had final responsibility for the decision to submit for publication.

## Acknowledgements

We thank Drs. Andrea Ciaranello and Sandra Nelson for their feedback on an early version of the manuscript, and Robert Jerris, PhD, Mark Gonzalez, PhD, Beverly Rogers, MD, and Yun F (Wayne) Wang, PhD and their laboratory staff at Children’s Healthcare of Atlanta and Grady Memorial Hospital for their contributions to clinical testing for SARS-CoV-2.

