## Supplementary Material for "Predictive value of isolated symptoms for diagnosis of SARS-CoV-2 infection in children tested during peak circulation of the delta variant"

**Supplementary Table 1**. Percentage of Unvaccinated Children Who Tested Positive by Symptom Status

| | *All Participants* | | *Excluding Participants with Fever* | |
|---|---|---|---|---|
| **Symptom Status** | **Total Participants** | **Participants Who Tested Positive (%)** | **Total Participants** | **Participants Who Tested Positive (%)** |
| 0 symptoms | 114 | 10 (8.8%) | 114 | 10 (8.8%) |
| 1 symptom | 71 | 23 (32.4%) | 58 | 15 (27.8%) |
| 2+ symptoms | 329 | 94 (28.6%) | 137 | 29 (21.2%) |

**Supplementary Table 2.** Percentage of Unvaccinated Children Who Had a Known or Suspected Close Contact and Who Tested Positive by Symptom Status

| **Symptom Status** | **Total Participants** | **Participants Who Had a Known or Suspected Close Contact (%)** | **Test Positive Rate (%) in Participants with a Known or Suspected Close Contact (%)** | **Participants Who Did Not Have a Known or Suspected Close Contact (%)** | **Test Positive Rate (%) in Participants with No Known or Suspected Close Contact** |
|---|---|---|---|---|---|
| *Unvaccinated participants* | | | | | |
| 0 symptom | 114 | 62 (54.4%) | 9 (14.5%) | 52 (45.6%) | 1 (1.9%) |
| 1 symptom | 71 | 41 (57.8%) | 19 (46.3%) | 30 (42.2%) | 4 (13.3%) |
| 2+ symptoms | 329 | 147 (44.7%) | 59 (40.1%) | 182 (55.3%) | 35 (19.2%) |

**Supplementary Table 3.** Diagnostic Accuracy of Isolated Symptoms in Unvaccinated Children who Presented with One Symptom

| Symptom | Overall N=71 | Negative N=48 | Positive N=23 | Sensitivity | Specificity | PPV | NPV |
|---|---|---|---|---|---|---|---|
| Fever | 13 | 5 | 8 | 0.35 (0.15, 0.54) | 0.90 (0.81, 0.98) | 0.62 (0.35, 0.88) | 0.74 (0.63, 0.85) |
| Chills | 0 | 0 | 0 | NA | NA | NA | NA |
| Congestion/ rhinorrhea | 18 | 16 | 2 | 0.09 (0.00, 0.20) | 0.67 (0.53, 0.80) | 0.11 (0.00, 0.26) | 0.60 (0.47, 0.74) |
| Cough | 18 | 12 | 6 | 0.26 (0.08, 0.44) | 0.75 (0.63, 0.87) | 0.33 (0.12, 0.55) | 0.68 (0.55, 0.80) |
| Headache | 5 | 3 | 2 | 0.09 (0.00, 0.20) | 0.94 (0.87, 1.00) | 0.40 (0.00, 0.83) | 0.68 (0.57, 0.79) |
| Sore throat | 9 | 4 | 5 | 0.22 (0.05, 0.39) | 0.92 (0.84, 0.99) | 0.56 (0.23, 0.88) | 0.71 (0.60, 0.82) |
| Fatigue | 0 | 0 | 0 | NA | NA | NA | NA |
| Arthralgias | 0 | 0 | 0 | NA | NA | NA | NA |
| Myalgias | 0 | 0 | 0 | NA | NA | NA | NA |
| Photophobia | 0 | 0 | 0 | NA | NA | NA | NA |
| Vomiting | 2 | 2 | 0 | 0.00 (0.00, 0.00) | 0.96 (0.90, 1.00) | 0.00 (0.00, 0.00) | 0.67 (0.56, 0.78) |
| Nausea | 0 | 0 | 0 | NA | NA | NA | NA |
| Diarrhea | 3 | 3 | 0 | 0.00 (0.00, 0.00) | 0.94 (0.87, 1.00) | 0.00 (0.00, 0.00) | 0.66 (0.55, 0.77) |
| Abdominal pain | 3 | 3 | 0 | 0.00 (0.00, 0.00) | 0.94 (0.87, 1.00) | 0.00 (0.00, 0.00) | 0.66 (0.55, 0.77) |
| Loss of sense of taste or smell | 0 | 0 | 0 | NA | NA | NA | NA |
| Shortness of breath | 0 | 0 | 0 | NA | NA | NA | NA |
